# Ozempic, Semaglutide Injection Misuse Reported Among a Nationally Representative Sample in the United States

**DOI:** 10.64898/2025.12.25.25343021

**Authors:** Orrin D. Ware

## Abstract

Ozempic, along with exercise and diet, is effective in helping individuals lose weight. Prior studies have reported misuse of Ozempic. This descriptive study used a nationally representative sample of persons in the U.S. from the National Survey on Drug Use and Health to identify self-reported use of an Ozempic injection for which the person was not prescribed. In 2024, 28,691 individuals ages 12 and older self-reported ever injecting Ozempic that was not prescribed to them. The entire sample who self-reported misusing Ozempic consisted of women aged at least 18 years (100%). Among this sample, more than 90% reported that their most recent misuse of Ozempic occurred in the past 12 months. Most of the sample was between the ages of 26 and 34 (64%), had a body mass index consistent with obesity (89%), was employed full-time (81%), was a college graduate or higher (91%), and had private health insurance coverage (81%). The entire sample reported consuming alcohol in the past 12 months (100%), and slightly more than half met the criteria for an alcohol use disorder diagnosis (53%). This is the first known study to characterize a nationally representative sample of persons who self-reported misusing Ozempic injections. Future studies are needed to examine the motivations for Ozempic misuse and to determine whether some individuals experience barriers to accessing it.

## INTRODUCTION

Ozempic (semaglutide) injections are a Food and Drug Administration-approved treatment for type 2 diabetes. Research has also shown that Ozempic induces significant weight loss even among individuals without type 2 diabetes.^1^ The effectiveness of Ozempic has increased its popularity as a weight loss drug, especially since a large proportion of the United States (U.S.) adult population has an obese body mass index. Approximately 40% of adults in the United States are obese, which can increase the risk of experiencing or exacerbating health-related problems.^2^ Overall, alongside diet and exercise, Ozempic is recognized as a public health intervention that can improve health outcomes for several individuals by increasing their ability to maintain a healthy weight.

Prescribed medications such as Ozempic carry the risk of misuse, which occurs when medications are used in ways they were not prescribed or by a person to whom they were not prescribed. Several posts expressing interest in anti-obesity medicines, such as Ozempic, have been expressed by social media users across various platforms,^3,4^ as have social media posts involving forged prescriptions for semaglutide.^5^ A study examining the Food and Drug Administration’s Adverse Events Reporting System found that use without prescriptions is more likely with semaglutide than with other glucagon-like peptide-1 receptor agonists (GLP1-RAs), such as tirzepatide.^5^ A study based in France found that approximately 2.2% users of GLP1-RAs such as Ozempic misuse the medications.^6^ Although the risk of misuse with Ozempic injections has been identified,^4,7^ there is a gap in the research literature regarding Ozempic injection misuse among a national sample in the U.S. Therefore, this descriptive study used a nationally representative sample of persons in the U.S. to identify self-reported use of an Ozempic injection for which the person was not prescribed.

## METHODS

The Substance Abuse and Mental Health Services Administration’s nationally representative National Survey on Drug Use and Health 2024, which is a health-based survey conducted at the household level, was used to identify individuals who ever injected Ozempic when it was not prescribed for them.^8^ Respondents are asked a series of questions regarding whether they have ever injected specific substances such as heroin and methamphetamine. Then, respondents are asked “Have you ever, even once, used a needle to inject any other drug/any drug that was not prescribed for you?”^8^ A yes response then permits these individuals to describe the specific other drugs that they injected. For this study, individuals were selected if they had the following response “Ozempic, Semaglutide injection”.

After selecting cases that reported ever injecting an “Ozempic, Semaglutide injection”, the analytic sampling weights were applied to identify the data on a population level. Descriptive statistics such as counts and percentages were used to characterize this sample based on the size of the area in which they currently reside (large, small, or non metropolitan areas), age, race and ethnicity, sex, body mass index (BMI), and binary responses (Yes/No) to whether they used other substances in the past 12 months (cigarettes, nicotine vaping, alcohol, cannabis, cocaine or crack cocaine, heroin, hallucinogens, inhalants, methamphetamine, tranquilizers, sedatives, and benzodiazepines). The time since the respondents’ last Ozempic injection misuse was also assessed: within the past 30 days, more than 30 days ago but within the past 12 months, and more than 12 months ago. Further, where the respondent obtained the needle they last used was also examined. After selecting the analytic sample, it was identified that all respondents were at least 18 years old. Therefore, additional variables were included to characterize the analytic sample. Employment status (employed full-time, employed part-time, unemployed) and whether the respondent was covered by private insurance, Medicare, and Medicaid/ Children’s Health Insurance Program (CHIP) were also assessed. Educational level was also examined, such as less than high school, high school graduate, some college/Associate’s degree, and college graduate. All study analyses and tables created to examine the sample of individuals who reported ever misusing an “Ozempic, Semaglutide injection” were done using SPSS Version 31.^9^ This study used de-identified publicly available data and was considered non-human subjects research by the University of North Carolina at Chapel Hill Institutional Review Board.

## RESULTS

After applying the population sample weights, a total of N = 28,691 (< 0.0% of the U.S. population) individuals in 2024, ages 12 and older, self-reported ever using an “Ozempic, Semaglutide injection” that was not prescribed to them. Characteristics and past-year substance use of this sample may be found in Table 1. Slightly more than half of the sample, at 53.4% lived in a small metropolitan area (n = 15,325). A little more than three in five individuals in the sample were aged 26 to 34 years (n = 18,340; 63.9%). Nearly nine in ten individuals in this sample had a body mass index consistent with obesity (n = 25,384; 88.5%). One hundred percent of the sample was female.

**Table 1.** Characteristics of a Nationally Representative Sample of Individuals in the United States who Self-Reported Ever Injecting Ozempic, Semaglutide that was not prescribed for them; N = 28,691.

| Variable | Count | Percentage |
| --- | --- | --- |
| **Area of Residence Size** |  |  |
| Large Metropolitan Area | 13,367 | 46.6% |
| Small Metropolitan Area | 15,325 | 53.4% |
| Nonmetropolitan Area | 0 | 0.0% |
| **Age** |  |  |
| 12-17 Years Old | 0 | 0.0% |
| 18-25 Years Old | 8,420 | 29.3% |
| 26-34 Years Old | 18,340 | 63.9% |
| 35-49 Years Old | 1,931 | 6.7% |
| 50-64 Years Old | 0 | 0.0% |
| 65 or Older | 0 | 0.0% |
| **Body Mass Index (BMI)** |  |  |
| Underweight | 0 | 0.0% |
| Normal Weight | 2,559 | 8.9% |
| Overweight | 748 | 2.6% |
| Obese | 25,384 | 88.5% |
| **Race and Ethnicity** |  |  |
| White | 9,093 | 31.7% |
| Black / African American | 6,085 | 21.2% |
| Native American / Alaska Native | 0 | 0.0% |
| Native Hawaiian Other Pacific Islander | 0 | 0.0% |
| Asian | 748 | 2.6% |
| More than One Race | 0 | 0.0% |
| Hispanic Any Race | 12,765 | 44.5% |
| **Sex** |  |  |
| Male | 0 | 0.0% |
| Female | 28,691 | 100.0% |
| **Past 12-Month Substances Used** |  |  |
| Cigarettes | 16,508 | 57.5% |
| Nicotine Vaping | 15,325 | 53.4% |
| Alcohol | 28,691 | 100.0% |
| Cannabis | 3,307 | 11.5% |
| Cocaine or Crack | 0 | 0.0% |
| Heroin | 0 | 0.0% |
| Hallucinogens | 0 | 0.0% |
| Inhalants | 0 | 0.0% |
| Methamphetamine | 0 | 0.0% |
| Tranquilizers | 5,575 | 19.4% |
| Sedatives | 0 | 0.0% |
| Benzodiazepines | 0 | 0.0% |

231 Some counts and percentages may be altered due to weighting and rounding error.

Approximately four in five individuals were employed full-time (n = 23,116; 80.6%), whereas one in five individuals was employed part-time (n = 5,575; 19.4%). Regarding insurance, 80.6% (n = 23,116) had private health insurance coverage, 0% (n = 0) had Medicare coverage, and 21.2% (n = 6,085) had Medicaid/Children’s Health Insurance Program (CHIP) coverage. A little more than nine in ten persons in the sample were college graduates or higher (n = 26,132; 91.1%), and nearly one in ten completed some college or had an Associate’s degree (n = 2,559; 8.9%). The majority of the sample, at 85.2% (n = 24,439), last misused an Ozempic injection more than 30 days ago but within the past 12 months, and 6% (n = 1,693) of the sample last misused an Ozempic injection within the last 30 days. Regarding where they obtained the last needle they used when injecting, approximately 68.0% (n = 19,523) bought it from a pharmacy, 18.7% (n = 5,351) obtained it from a drug dealer or it came with the drugs, and 4.4% (n =1,258) bought the needle online/from the internet.

As shown in Table 1, 57.5% (n = 16,508) of the sample smoked cigarettes in the past 12 months, and 53.4% (n = 15,325) vaped nicotine in the past 12 months. The entire sample, 100% (n = 28,691), drank alcohol in the past 12 months. After identifying the high prevalence of past-year alcohol consumption in this sample, the prevalence of past-year alcohol use disorder was also examined. Slightly more than half of the sample met the criteria for an alcohol use disorder at 53.4% (n = 15,325), whereas 46.6% (n = 13,367) did not.

## DISCUSSION

Considering evidence suggesting the potential for Ozempic to be misused,^6^ this study describes a nationally representative sample of persons reporting injecting the medication that was not prescribed to them. Overall, the number of individuals reporting that they had injected Ozempic without a prescription in this nationally representative study was 28,691, representing less than 0.0% of the U.S. population. Furthermore, slightly more than 90% of this sample reported that they last misused an Ozempic injection within the past 12 months. Characteristics of this sample were also identified in this study.

All the respondents were Female. A systematic review on patient experiences with GLP1-RAs found that females may report more benefits of using these medications than males.^10^ Most of the respondents lived in a small metropolitan area, were between ages 26 and 34, were employed full-time, and were covered by private health insurance. Given the weight-management properties of Ozempic, which likely contributed to its misuse in the sample, nearly 90% of participants had a body mass index consistent with obesity. Regarding past-year substance use, 100% of the sample reported consuming alcohol, and 53% met the criteria for an alcohol use disorder. The prevalence of alcohol consumption among this sample should be considered alongside findings that semaglutide may decrease overall alcohol consumption and alcohol craving among some persons.^11,12^ Future studies are needed to determine whether individuals misusing Ozempic do so for weight loss purposes, to reduce alcohol consumption/craving, or a mixture of both. Nicotine vaping and cigarette smoking were reported by over half of the sample at 53% and 58% respectively.

This study provides a sociodemographic snapshot of a previously undescribed sample. By providing this pilot data, which characterizes the sample, future studies may collect data on motivations for and frequency of Ozempic misuse. Furthermore, future studies focusing specifically on individuals who misuse Ozempic may determine whether the characteristics of respondents in their sample align with the pilot results presented in this current study.

### Limitations

Data from the National Survey on Drug Use and Health can be affected by participation and response bias, particularly because some individuals may be less willing to report misusing prescribed medications. This study used body mass index (BMI) scores to characterize the sample; however, BMI does not account for muscle mass. Another limitation is that respondents were not asked directly whether they have ever used an “Ozempic, Semaglutide injection” that was not prescribed to them. Instead, they were asked about other drugs they had ever injected that were not prescribed to them, and they could provide a binary Yes/No response and then mention “Ozempic, Semaglutide injection” as one of the substances. There would likely be a higher prevalence of persons self-reporting “Ozempic, Semaglutide injection” misuse if they were asked directly. Therefore, while this pilot study identifies respondents who self-report misusing “Ozempic, Semaglutide injection,” it does not provide a true prevalence because respondents were not directly asked.

## CONCLUSION

In 2024, 28,691 individuals self-reported ever injecting Ozempic that was not prescribed to them. The entire sample who self-reported misusing Ozempic consisted of women aged at least 18 years old who drank alcohol in the past 12 months (100%). Nearly 90% of the sample had a body mass index that was consistent with obesity, and slightly more than half met the criteria for an alcohol use disorder diagnosis (53%). This is the first known study to characterize a nationally representative sample of persons who self-reported misusing Ozempic injections. Future studies are needed to examine the motivations for Ozempic misuse and to determine whether some individuals experience barriers to accessing it.

## Data Availability

The Substance Abuse and Mental Health Services Administration nationally representative National Survey on Drug Use and Health 2024: https://www.samhsa.gov/data/data-we-collect/nsduh-national-survey-drug-use-and-health.

